# ALDH2/eIF3E Interaction Modulates Protein Translation Critical for Cardiomyocyte Ferroptosis in Acute Myocardial Ischemia Injury

**DOI:** 10.1101/2024.08.19.24312276

**Authors:** Xin Chen, Xiujian Yu, Xiaodong Xu, Rui Li, Ningning Liang, Lili Zhang, Luxiao Li, Jingyu Zhang, Mingyao Zhou, Tongwei Lv, Haoran Ma, Yongqiang Wang, Yanwen Ye, Chunzhao Yin, Shiting Chen, Hui Huang, Jinwei Tian, Aijun Sun, Weiyuan Wang, Dewen Yan, Pan Li, Huangtian Yang, Shanshan Zhong, Huiyong Yin

**Affiliations:** CAS Key Laboratory of Nutrition, Metabolism and Food Safety, Shanghai Institute of Nutrition and Health (SINH), University of Chinese Academy of Sciences (UCAS), Chinese Academy of Sciences (CAS), Shanghai, China 200031; CAS Key Laboratory of Tissue Microenvironment and Tumor, Laboratory of Molecular Cardiology, SINH, UCAS, Shanghai, China 200031; School of Life Science and Technology, ShanghaiTech University, Shanghai, China; Department of Biomedical Sciences, Jockey Club College of Veterinary Medicine and Life Sciences, Tung Biomedical Science Center, State Key Laboratory of Marine Pollution (SKLMP), The Shenzhen Research Institute and Futian Research Institute, City University of Hong Kong, Hong Kong, China; Department of Cardiology, Changhai Hospital, Naval Medical University, Shanghai, China; Bio-med Big Data Center, CAS Key Laboratory of Computational Biology, CAS Center for Excellence in Molecular Cell Science, SINH, UCAS, Shanghai, China 200031; Department of Cardiology, The Eighth Affiliated Hospital, Joint Laboratory of Guangdong-Hong Kong-Macao Universities for Nutritional Metabolism and Precise Prevention and Control of Major Chronic Diseases, Sun Yat-sen University, Shenzhen, China; Department of Cardiology, Second Affiliated Hospital of Harbin Medical University, Key Laboratory of Myocardial Ischemia, Chinese Ministry of Education, Harbin, China; Department of Cardiology, Zhongshan Hospital, Fudan University, Shanghai Institute of Cardiovascular Diseases, 180 Fenglin Road, Shanghai 200032, China; Institutes of Biomedical Sciences, Fudan University, 131 Dongan Road, Shanghai 200032, China; Department of Endocrinology, Shenzhen Second People’s Hospital, the First Affiliated Hospital of Shenzhen University, Health Science Center of Shenzhen University, Shenzhen Clinical Research Center for Metabolic Diseases, Shenzhen Center for Diabetes Control and Prevention, No. 3002, Sungang West Road, Futian District, Shenzhen, 518035, Guangdong Province, China

**Keywords:** ALDH2, myocardial infarction, ferroptosis, protein translation

## Abstract

**BACKGROUND:** As an iron-dependent form of regulated cell death caused by lipid peroxidation, ferroptosis has been implicated in ischemic injury but the underlying mechanisms in acute myocardial infarction (AMI) remain poorly defined. Acetaldehyde dehydrogenase 2 (ALDH2) catalyzes detoxification of lipid aldehydes derived from lipid peroxidation and acetaldehydes from alcohol consumption. The Glu504Lys polymorphism of ALDH2 (rs671, ALDH2 *2), affecting around 8% world population and 40% East Asians, is associated with increased risk of MI. This study aims to investigate the role of ALDH2 and ferroptosis in MI.

**METHODS:** A Chinese cohort of 177 acute heart failure patients with ALDH2 wild type and ALDH2 *2 were enrolled. MI mouse model of left anterior descending coronary artery ligation (LAD) was conducted on wild type, ALDH2 *2, and mice with cardiomyocyte-specific knock down of eukaryotic translation initiation factor 3 subunit E (eIF3E) by adeno-associated virus. The lipid peroxidation products were measured by mass spectrometry-based lipidomics and metabolomics in human plasma and mouse serum samples as well as in mouse heart tissues.

**RESULTS:** Human ALDH2 *2 carriers exhibit more severe heart failure post-AMI with features of ferroptosis in blood samples through lipidomic analysis, including increased levels of multiple classes of oxidized phospholipids, and decreased levels of antioxidants, such as Coenzyme Q-10 (Co-Q10) and tetrahydrobiopterin (BH4). Similar features were observed in MI mouse models of ALDH2 *2, whereas ferroptosis inhibition by Fer-1 significantly improved heart functions and reversed ferroptosis markers. Importantly, ALDH2 *2 led to significantly decreased protein levels of ALDH2, whereas ferroptosis related proteins including Transferrin receptor (TFRC), Acyl-CoA synthetase long chain family member 4 (ACSL4), and Heme oxygenase 1 (HMOX1) were upregulated specifically in the infarct heart tissues. Mechanistically, ALDH2 physically interacted with eIF3E to modulate translation of critical proteins involved in ferroptosis, and ALDH2 deficiency in ALDH2 *2 mutant predisposes cardiomyocytes to ferroptosis by promoting Tfrc/Acsl4/Hmox1 translation. Consistently, cardiomyocytes-specific eIF3E knock down restored ALDH2 *2 cardiac function by attenuating ferroptosis in MI.

**CONCLUSIONS:** ALDH2 *2 aggravates acute heart failure in MI through promoting cardiomyocytes ferroptosis, and targeting ferroptosis may be a potential therapeutic target for treating AMI, especially for ALDH2 *2 carriers.

**Clinical Perspective:** *What Is New?:*

- ALDH2 *2 mutant aggravates acute heart failure through promoting cardiomyocytes ferroptosis after myocardial infarction.
- Active translation of proteins critical for ferroptosis predisposes cardiomyocytes of ALDH2 deficiency to ferroptosis through attenuated interactions with elF3E and ALDH2.
- Targeting ferroptosis induced by ALDH2 deficiency rescued heart functions post-AMI.

*What Are the Clinical Implications?:*

- Circulating oxidized phospholipids and other features associated with ferroptosis may serve as markers for AMI, especially for ALDH2 *2 carriers.
- Inhibiting ferroptosis is a viable cardioprotective therapeutic strategy for MI injury particularly for ALDH2 *2 carriers.

## INTRODUCTION

Acute myocardial infarction (AMI) is a leading cause of deaths worldwide, and ischemia heart injury remains the major contributor to heart failure post-AMI.^1–4^ Mounting evidence has implicated various forms of cardiomyocyte cell death in ischemia injury, such as apoptosis, necroptosis, pyroptosis, and ferroptosis.^5, 6^ Each form of cell death plays an important role in the context of ischemia injury and specific targeting different cell death may have enormous impact on developing therapeutic strategies for MI.^7^

Ferroptosis is a newly discovered form of regulated cell death primarily caused by lipid peroxidation and dysregulation of iron metabolism.^8–10^ Multiple reactive oxygen species, generated from Fenton chemistry (ferric ion and hydrogen peroxide), lipoxygenases (LOX), cytochrome P450 oxidoreductase (POR), or NADPH oxidases (NOX), attack polyunsaturated fatty acids (PUFA) in cellular membranes, and oxidized lipids have been considered hallmarks of lipid peroxidation and ferroptosis.^11, 12^ On the other hand, cellular antioxidant systems, including the glutathione peroxidase 4 (GPX4), Nrf2/KEAP1, ubiquinol recycling (ferroptosis suppressor protein 1-coenzyme Q10, Co-Q10), and GTP cyclohydrolase 1-mediated tetrahydrobiopterin (BH4) production, are critical for counteracting lipid peroxidation, which proceeds ferroptosis.^8^ Acyl-CoA synthetase long-chain family member 4 (ACSL4) plays an important role in synthesis PUFAs for the cellular membranes during ferroptosis.^13^ ACSL4 also preferentially activates arachidonate and eicosapentaenoate as substrates and modulates prostaglandin E2 secretion.^14^ Mounting evidence for the past decade has implicated ferroptosis in various human diseases, especially ischaemia–reperfusion injury; however, the underlying mechanisms of ferroptosis in AMI remain poorly defined.^9^

Aldehyde dehydrogenase 2 (ALDH2) is a critical enzyme catalyzing oxidation of acetaldehydes from alcohol or endogenous lipid aldehydes from lipid peroxidation.^15^ A loss-of-function single-nucleotide polymorphism (SNP) in the ALDH2 gene, ALDH2 *2 (or ALDH2 rs671), affecting up to 8% of the global population and around 30-50% of the East Asian population, has been associated with increased risk of MI and other cardiovascular diseases (CVD).^16–23^ Most previous studies have attributed the increased risks of CVD in ALDH2*2 carriers to the accumulation of aldehydes as a result of significantly decreased enzymatic function of ALDH2.^24, 25^ We and others have discovered multiple non-enzymatic functions of ALDH2 in the pathogenesis of CVD.^17, 26–28^ However, it remains elusive whether ALDH2 regulates lipid peroxidation and ferroptosis in MI.^17, 29, 30^

In this study, we discovered a novel mechanism by which ALDH2 physically interacts with eukaryotic translation initiation factor 3 subunit E (eIF3E) to modulate the translation of several key proteins involved in ferroptosis, whereas protein deficiency in ALDH2 *2 promotes cardiomyocyte ferroptosis through enhancing translation of Tfrc/Acsl4/Hmox1. On the other hand, knockdown eIF3E or inhibiting ferroptosis by Fer-1 attenuates heart failure in ALDH2 *2 mice, suggesting inhibiting ferroptosis as a novel therapeutic target for ischemia heart injury.

## METHODS

The authors declare that all supporting data are available within the article and its online supplementary files. The data, all protocols, and study materials are available to other researchers for purposes of reproducing the results. The detailed methods used in this study are provided in the online-only **Supplemental Material**.

### Myocardial Infarction Injury Model and Study Designs

All protocols involving animals were performed in compliance with relevant ethical guidelines. The experimental design was approved by the Animal Care and Use Committee of the Shanghai Institute of Nutrition and Health, University of Chinese Academy of Sciences (CAS). Experimental mice were housed in a pathogen-free, temperature- and light-controlled animal facility under a 12h light/dark cycle. Male C57BL/6 and ALDH2 *2 mutant mice at 10 weeks-old were anesthetized via intraperitoneal injection (i.p.) of 50 mg/kg sodium pentobarbital and mechanically ventilated using a volume regulated respirator (SAR830, Cwe Incorporated). Body temperature was maintained at 37°C throughout the surgical procedure. The mice were randomly selected for the sham group and the induction of MI. Ischemia was produced through ligation of the left anterior descending (LAD) coronary artery with 7-0 prolene suture, while sham animals underwent an identical procedure with the exception that tension was not placed on the suture ends. The skin was then closed over the exteriorized suture ends with 5-0 prolene suture, and the mice were returned to the rearing cage for continued feeding. Vehicle (1% DMSO in 0.9% saline i.p.) or Fer-1 (10 mg/kg in 0.9% saline i.p.) was administered 24 and 2 hours before ischemia.

### Human Peripheral Blood Samples and Coronary Artery Tissues

Patients diagnosed with ST-segment–elevation myocardial infarction were recruited from the Changhai Hospital of Shanghai. The plasma samples of patients were collected before surgery and total DNA were isolated from peripheral blood plasma. The left ventricular tissues were obtained from 3 patients with dilated cardiomyopathy with heart failure during heart transplantation at the cardiac surgery department of Changhai Hospital. The study protocol was approved by the institutional ethics committee of Changhai Hospital of Shanghai on research in humans. Written informed consent was obtained from each patient included in the study.

### Analysis of Oxidized Peroxides

400 μL chloroform and 200 μL methanol (HPLC grade, contained 0.005% Butylated hydroxytoluene) was added to 200 μL tissue homogenates or plasma. Upon thawing, 100 ng PE (D16:1) and 100 ng PC (D14:1) internal standard was added to each sample. The organic phase was dried under vacuum and resuspended in 50 μL of a mixture of 100% solvent B (methanol/ isopropanol 30:40, v/v, conclude Amide acetate). Samples (10 μL) were then injected into an LC MS system. Chromatography was performed using a HILIC HPLC column (Luna 5 μm, 100 Å, 50 × 2 mm, Phenomenex) at a flow rate of 0.350 mL/ min. Mass spectrometric analysis was performed in the negative ion mode using multiple-reaction monitoring (MRM) of specific precursor–product ion m/z transitions upon collision-induced dissociation. The precursor negative ions monitored were the molecular ions [M – H] − for PE, and the acetate adducts [M +CH3COO] − for PC. Identity was further verified by monitoring at the same time, using polarity switching, the positive molecular ions [M + H] + for both PC and PE molecular species. The specific precursor–product pairs monitored in negative-ion mode and used for quantification were used as described.^31, 32^ The specific precursor–product pairs monitored in negative-ion mode and used for quantification were showed in the Table S4. Results were reported as the ratio of the integrated area of each analyte and the integrated area of the corresponding internal standard, and then multiplied to the concentration of corresponding internal standard.

### Statistical Analysis

All mice were assigned to randomized groups before experiments. And experiments including cell, animal and human experiments were analyzed using two-tailed Student t test for normal data and the Mann-Whitney test for unpaired data by GraphPad Prism 9.0 software. For comparisons between multiple groups, a one-way or 2-way ANOVA was performed, followed by the Tukey multiple comparisons post-test. All data were reported as the means ± SEM and the means ± SD from at least three independent experiments. Differences were considered statistically significant at *P* < 0.05 and statistics significance was shown as \**P* < 0.05; \*\**P* < 0.01; \*\*\**P* < 0.001.

## RESULTS

### ALDH2 *2 is Positively Associated with Severity of Acute Heart Failure post-MI

To explore the relationship between ALDH2 *2 and acute heart failure (AHF) induced by myocardial infarction (MI), we enrolled a cohort of 177 patients with ALDH2 *1 (wild type, WT, n=111) and ALDH2 variant (ALDH2 *2, n=66), respectively. The clinical characteristics of all these patients were summarized in Table 1. The white blood cells were used for ALDH2 genotyping and plasma samples were used for metabolomics and lipidomics analysis (Figure 1A). The major known risk factors for HF between these two groups were similar, such as age, sex, BMI, levels of blood lipids, family history of CVD, and metabolic comorbidities including hypertension and diabetes mellitus. We used levels of brain natriuretic peptide (BNP) as the proven prognostic factor for patients with acute decompensated heart failure (ADHF) and BNP greater than 100pg/mL is defined as acute HF.^33, 34^ The patients carrying ALDH2 *2 variant had much higher levels of BNP compared to WT. Consistently, levels of other clinical parameters for AHF, such as LDH and CRP, were also much higher than those in WT. On the other hand, the heart functions deteriorated more in ALDH2 variant than WT with significantly decreased ejection fraction (EF) and fraction shortening (FS) indices, increased left ventricular end-systolic dimension (LVESD) and left ventricular end-diastolic volume (LVEDV). Interestingly, prevalence of drinking was obviously lower in the ALDH2 *2 than WT, consistent with previous reports (Table 1).

**Table 1.**
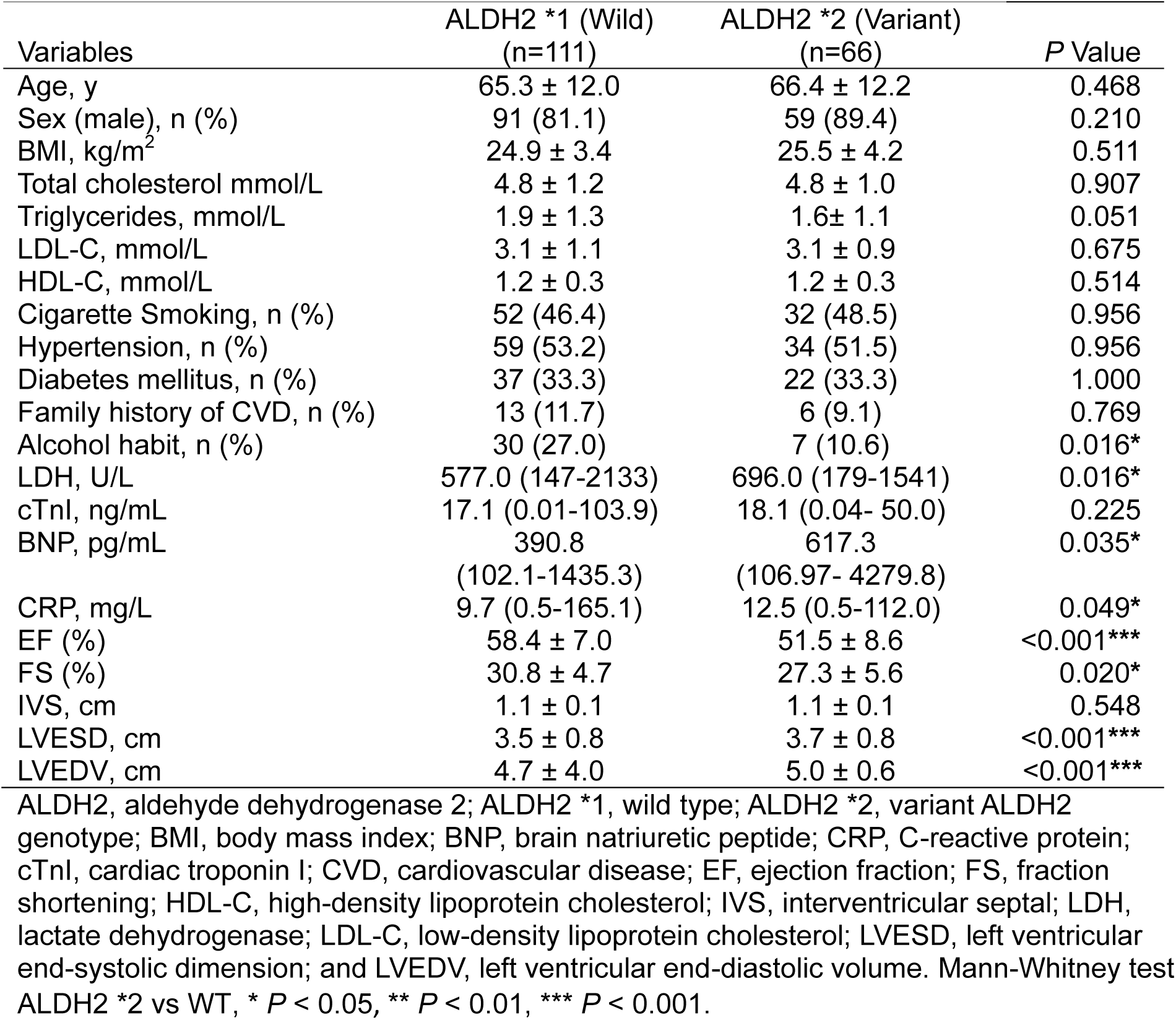
Clinical characteristics of human samples from AHF patients post-MI.

**Figure 1.**
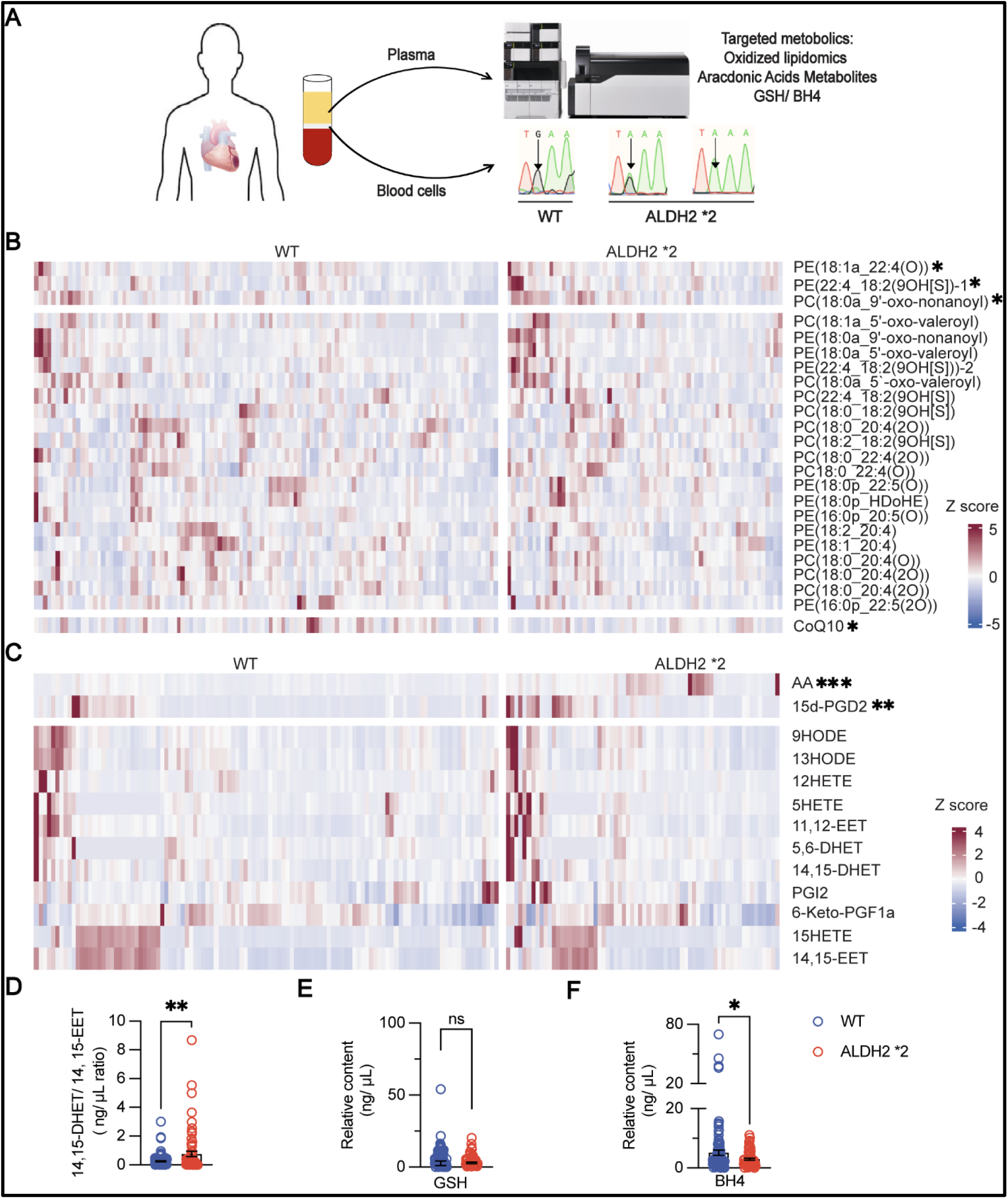
Targeted metabolomic analysis of oxidative lipids and its metabolites in the human plasma samples from ALDH2 WT and ALDH2 *2 carriers. **A**, experimental design. BNP greater than 100 pg/mL was classified as heart failure. WT, ALDH2 *2 mutation was verified by direct sequencing of PCR-amplified genomic DNA (WT, n= 111; ALDH2 *2, n= 66). **B**, relative levels of oxidized phospholipids and CoQ10 in the plasma of WT and ALDH2 *2 heart failure patients. **C**, relative levels of bioactive lipids in the plasma of WT and ALDH2 *2 patients derived from free arachidonic acid and linoleic acid. Samples were averaged and normalized to WT group. Each metabolite was normalized to the internal standards. **D**, 14,15-DHET/14,15-EET ratio increased in the plasma of ALDH2 *2 patients. **E** and **F**, GSH and BH4 relative content in the plasma of WT and ALDH2 *2 patients (WT, n= 111; ALDH2 *2, n=66). *P* > 0.05 means ns, \**P* < 0.05, \*\**P* < 0.01 and \*\*\**P* < 0.001 from Mann-Whitney test. Data are mean ± SEM. AA, arachidonic acid; BH4, tetrahydrobiopterin; BNP, brain natriuretic peptide; DHET, dihydroxyeicosatrienoic acids; EET, epoxyeicosatrienoic acids; GSH, glutathione; WT, wild type.

Taken together, patients with ALDH2 *2 variant manifest aggravated AHF post-MI accompanied with decreased alcohol intake compared to those with ALDH2 WT.

### Targeted Metabolomic Analysis Identified Features of Ferroptosis in The Plasma of ALDH2 *2 AHF Patients post-MI

ALDH2 is a key enzyme in detoxifying ethanol-derived acetaldehyde and endogenous lipid aldehydes generated from lipid peroxidation,^29, 35^ and the latter has been closely linked to ferroptosis. However, it remains elusive whether ferroptosis is involved in ALDH2 *2 to exacerbate acute heat failure post-MI. We performed targeted oxidative lipidomics in the plasma samples by using liquid chromatography-mass spectrometry methods (LC-MS) and found that several classes of oxidized lipids significantly increased in ALDH2 *2 patients compared to WT, such as oxidized phosphatidylcholine (PC), phosphatidylethanolamine (PE), and truncated side chain oxidation products of PC and PE (Figure 1A and 1B).

Next, we used a separate targeted metabolomic method to detect bioactive lipids derived from free arachidonic acid (AA) and linoleic acid. Interestingly, we found that 15d-PGD_2_, a downstream metabolite of prostaglandin D_2_ (PGD_2_) in the cyclooxygenase (COX) pathway, and ratios of two metabolites from cytochrome P450 pathways of arachidonic acid, and 14,15-DHET/14,15-EET, were significantly elevated in the plasma of ALDH2 *2 patients (Figure 1C and 1D), consistent with a previous study in which these parameters represented one of the strongest predictors of one-year death in HF patients.^36^ Moreover, the levels of two important antioxidants coenzyme Q10 (CoQ10) and tetrahydrobiopterin (BH4) both decreased, whereas no difference of the glutathione (GSH) was observed (Figure 1E and 1F). Noteworthy, the levels of CoQ10 and BH4 have been closely correlated with heart failure and ferroptosis pathway.^37–39^

Together, all these metabolic data with elevated levels of oxidized lipids but decreased levels of BH4 and CoQ10 strongly suggested the involvement of ferroptosis in AHF of patients carrying ALDH2 *2 variant.

### ALDH2 *2 Mice Showed Worse Heart Function in MI Model

To provide further evidence that ALDH2 mutant exacerbates acute heart failure post MI through ferroptosis pathways, we employed an MI mouse model by ligating the left anterior descending (LAD) coronary artery and measuring the heart function by echocardiography (Echo) at the indicated time points (Figure 2A). The heart functions deteriorated more in ALDH2 *2 mice compared to WT as indicated by lower LVEF and LVFS throughout the experiments (Figure 2B and 2C). After MI, the first 4 days are acute inflammation phase with intense infiltration of inflammatory cells, followed by resolution and repair with active resolution of inflammation, quiescence of cell activity, scar stabilization and maturation from 7-14 days in mice.^40, 41^ Then we set 3 days post MI as the early acute MI stage, the makers of heart injury, mRNA level of Myh7, Nppa and Nppb in the infarct tissues were also increased significantly in ALDH2 *2 mice at day 3 post MI (Figure 2D). The mitochondria damage markers mt 16s and mt ND4 in the serum, and number of leukocytes in the blood of ALDH2 *2 AMI mice all increased although the number of monocytes did not change (Figure S1A-E).

**Figure 2.**
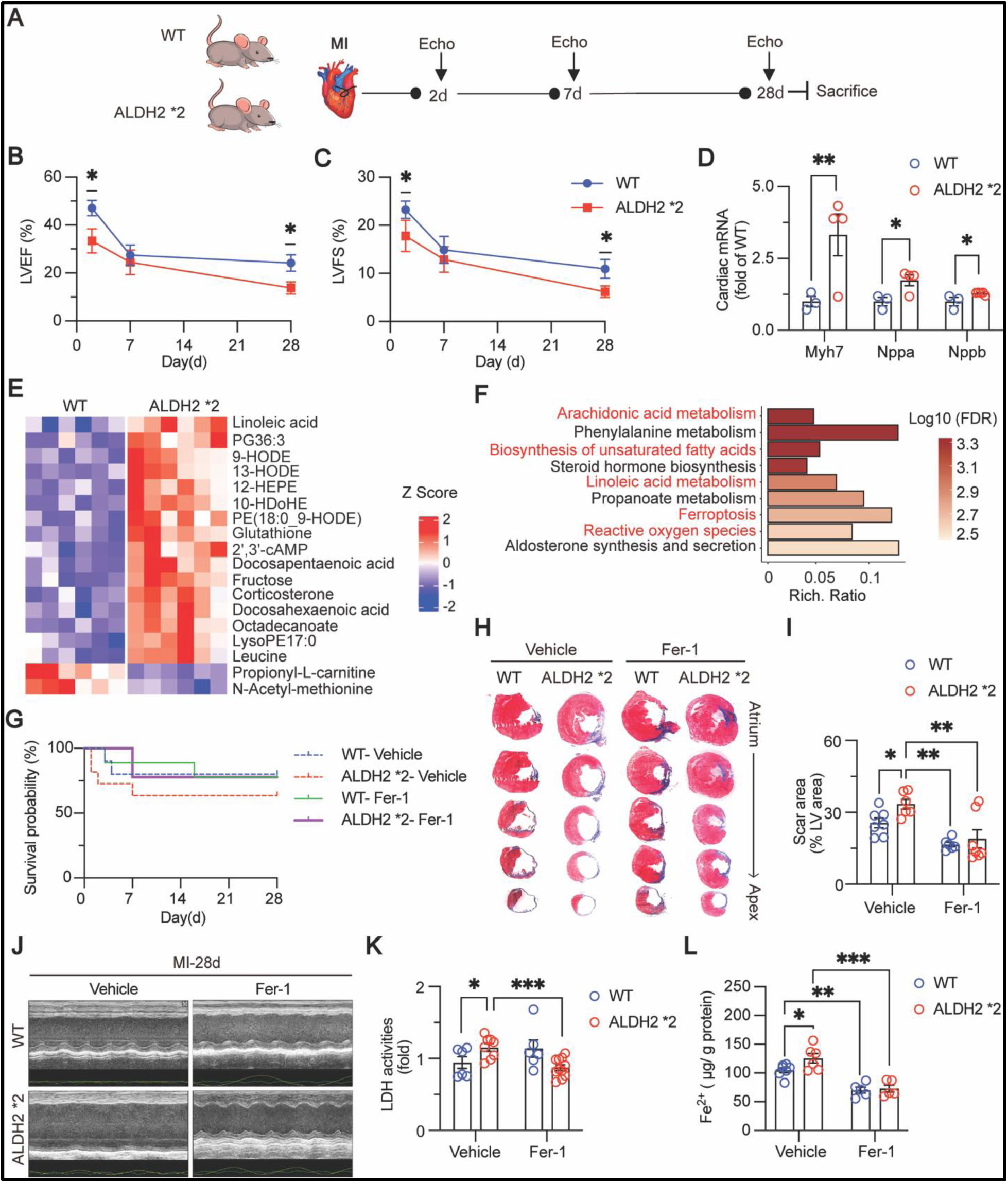
ALDH2 *2 exacerbated heart failure mainly through inducing ferroptosis in mice post-AMI. **A**, schematic diagram of the animal experiment. WT and ALDH2 *2 mice for surgery were at 10∼12 weeks old, and cardiac functions were measured at 2 days, 7 days, and 28 days post-AMI. **B** and **C,** quantification of echocardiography parameters (LVEF and LVFS, n=7, and 7, respectively). Results were reported as the mean ± SEM. \**P* < 0.05 from Student t test of WT control vs ALDH2 *2 mutant at the specified time point. **D**, mRNA of cardiac injury biomarkers Myh7, Nppa, Nppb in the infarct area post-AMI determined by RT-qPCR (n=3, 4 from each group). Results were reported as the mean ± SEM. \**P* < 0.05 from 2-way ANOVA test WT control vs ALDH2 *2 mutant. **E** and **F**, untargeted metabolomics results of AMI mice serum (n=6 from each group) by using LC MS. **E**, each row in the heatmap represented a specific metabolite that had significantly different expression levels in comparisons between WT and ALDH2 *2, the expression of which was normalized across the column, with high expression shown in red and low in blue. **F**, KEGG pathway analysis of untargeted metabolomics data from the serum of WT and ALDH2 *2 mice 3-day post-AMI. FC ≥ 2 and FDR < 0.05. **G**, 28 days survival rate of WT and ALDH2 *2 mice after MI with ferroptosis inhibitor Fer-1(1 mg·kg^-1^,24 and 2 hours i.p. before surgery) pretreatment or not (WT-Vehicle, ALDH2 *2-Vehicle, WT-Fer-1, ALDH2 *2-Fer-1, n=10, 11, 10, and 9, respectively). **H**, representative images of Masson Trichrome staining and **I**, quantitation of scar areas in WT and ALDH2 *2 mice hearts 28 days after surgery (n=7, 6, 6, and 7, respectively). Scale bar, 1mm. Results were reported as the mean ± SEM. \**P* < 0.05 and \*\**P* < 0.01 from Student t test WT vs ALDH2 *2 mutant with Fer-1 pretreatment or not. **J**, corresponding representative of echocardiographic images of **H** 28 days after MI. **K**, fold change of LDH activity in the serum 3-day post-AMI (WT vehicle, ALDH2 *2 vehicle, WT Fer-1, ALDH2 *2 Fer-1, n=6, 8, 6, and 11, respectively). Results were reported as the mean ± SEM. \**P* < 0.05, and \*\*\**P* < 0.001 from 2-way ANOVA test. **L**, the contents of Fe (II) in the infarct tissues of WT and ALDH2 *2 with vehicle or Fer-1 pretreatment 3-day post-AMI. Results were reported as the mean ± SEM. \**P* < 0.05, \*\**P* < 0.01, and \*\*\**P* < 0.001 from 2-way ANOVA test. Echo, echocardiography; FC, fold change; FDR, false discovery rate; Fer-1, ferrostatin-1; i.p. intraperitoneal injection; KEGG, kyoto encyclopedia of genes and genomes; LC MS, liquid chromatograph mass spectrometer; LDH, lactate dehydrogenase; LVEF, left ventricular ejection fraction; LVFS, left ventricular fraction shortening; MI, myocardial infarction; Myh7, myosin heavy chain 7; Nppa, natriuretic peptide A; Nppb, natriuretic peptide B.

All these data are consistent with the observations in patients in which ALDH2 *2 is linked to acute heart failure post MI.

### Ferroptosis was Involved in Acute Heart Failure in ALDH2 *2 Mice and Inhibition of Ferroptosis Improved Heart Function

We next performed untargeted metabolomics in the serum of mice post-AMI and found that levels of bioactive lipids derived from PUFAs, such as arachidonic acid, linoleic acid, and oxidized PE were all elevated in ALDH2 *2 group compared to WT (Figure 2E). The pathway enrichment analysis converged on ferroptosis and ROS (Figure 2F), suggesting that ferroptosis was involved in ALDH2 *2 to accelerate acute heart failure post MI.

To provide further evidence that ALDH2 *2 exacerbates heart injury through ferroptosis during AMI, we used a widely used ferroptosis inhibitor Ferrostatin-1 (Fer-1) to pretreat mice before MI surgery. The mice were randomized to the Fer-1 and control group based on LVEF, LVFS and body weights (Figure S2A and S2B). Interestingly, Fer-1 pretreatment helped mice survival (Figure 2G) and improved the heart functions of ALDH2 *2 post-MI as indicated by LVEF and LVFS (Figure S2C and S2D). Consistently, the heart injury shown as the scar areas exacerbated in ALDH2 *2 mice was also attenuated with Fer-1 pretreatment (Figure 2H and 2I). Echo results showed the heart ejection function was also improved with Fer-1 pretreatment (Figure 2J). Furthermore, the increased LDH activities in ALDH2 *2 at 3-day post-AMI were disappeared with Fer-1 pretreatment (Figure 2K). Lastly, the increased levels of Fe^2+^ in the infarct tissues of ALDH2 *2 mice was attenuated with Fer-1 pretreatment (Figure 2L), while the elevated levels of bioactive lipids in the serum and oxidized phospholipids in the infarct tissues of ALDH2 *2 mice were also significantly decreased (Figure S2E and S2F). All these data revealed that inhibiting ferroptosis in ALDH2 *2 at AMI stage confers benefits in protecting heart from MI injury and improving heart functions.

Taken together, ALDH2 *2 mutant exacerbated acute heart failure through ferroptosis post MI.

### Ferroptosis-related Protein TFRC, ACSL4, and HMOX1 Increased Specifically in ALDH2 *2 Ischemia Tissue

To further support the role of ferroptosis in cardiac injury of ALDH2 *2 mutant at post-AMI stage, we performed targeted lipidomic analysis on oxidized phospholipids, and found that cardiac infarct tissues had increased levels of multiple oxidized PC and PE, and these oxidized lipids were further elevated in MI of ALDH2 *2 mice compared to WT (Figure 3A). Further, we used inductively coupled plasma (ICP) MS to detect total iron and other metal ions in the infarct tissues and found total iron content increased in ALDH2 *2 (Figure 3B), whereas there was no difference between the levels of selenium. We also found Transferrin increased in the serum of ALDH2 *2 post-AMI (Figure 3C). Moreover, immunohistochemical analysis of heart slices staining that TFRC (transferrin receptor) was also significantly upregulated specifically in the infarct areas of ALDH2 *2 (Figure 3D).

**Figure 3.**
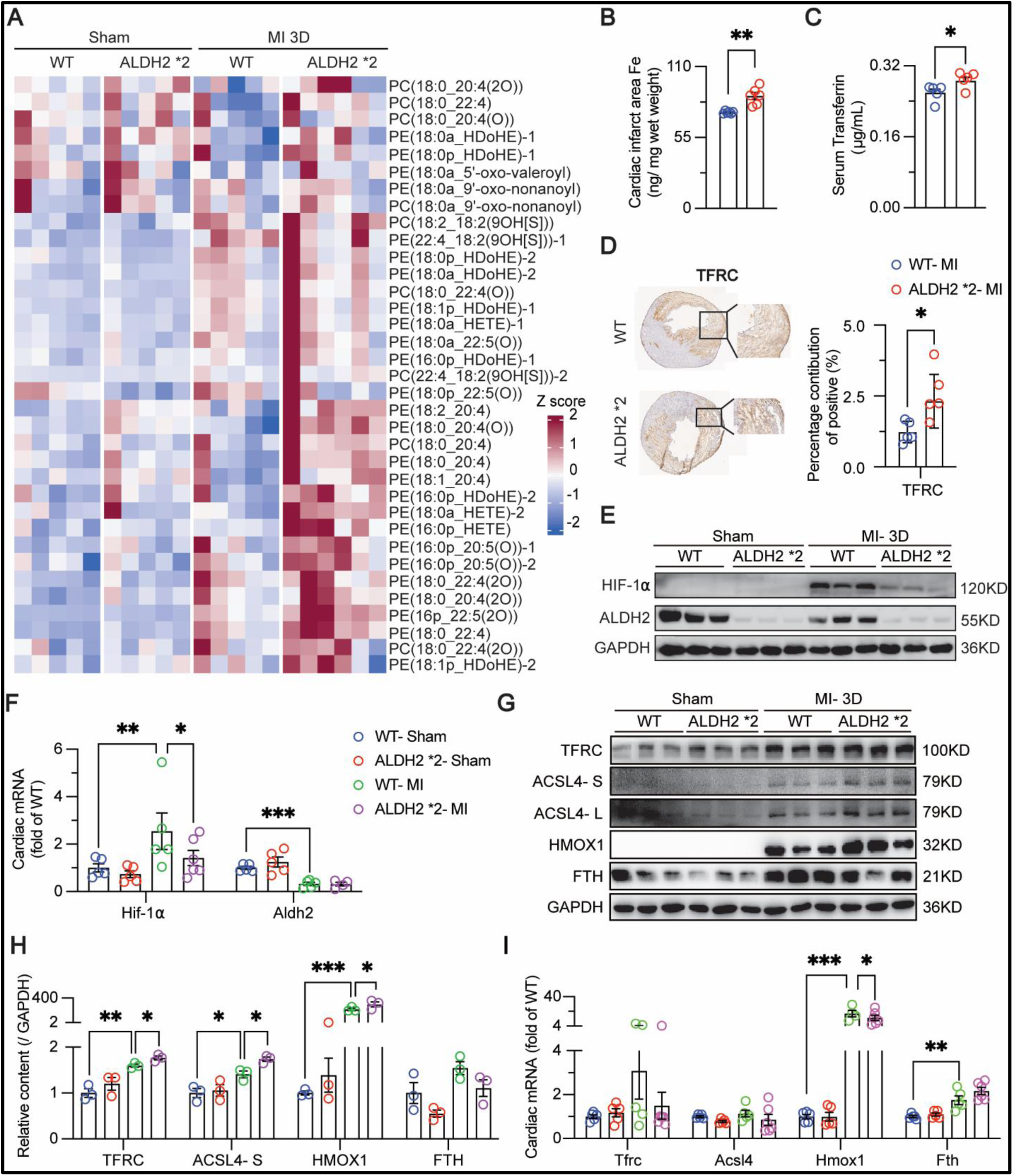
Oxidized phospholipids and ferroptosis-related proteins TFRC, ACSL4 and HMOX1 increased specifically in the infarct tissues of ALDH2 *2 mice. **A**, targeted metabolomic analysis of oxidized phospholipids in the infarct area of mouse heart (WT sham, ALDH2 *2 sham, WT MI 3d, and ALDH2 *2 MI 3d, n=5, 5, 5, and 6, respectively). **B**, total Fe content detected by ICP-MS in the cardiac infarct tissues (WT, ALDH2 *2, n=6, and 6, respectively). \*\**P* < 0.01 from Student t test of WT control vs ALDH2 *2 mutant. **C**, transferrin levels in the serum (n=5 for each group). \**P* < 0.05 from Student t test of WT control vs ALDH2 *2 mutant. **D**, representative immunohistochemistry images and quantitation of TFRC protein in the cardiac sections obtained from WT and ALDH2 *2 mice 3-day post-AMI (n=5 for each group). Scale bar, 1mm. \**P* < 0.05 from Student t test. **E**, representative immunoblotting images of HIF-1α and ALDH2 in the infarct tissues of WT and ALDH2 *2 3-day post-AMI or sham surgery (n=3 for each group). **F**, correlated mRNA level of Hif-1α and Aldh2 in WT and ALDH2 *2 mice with sham or MI pretreatment (WT-Sham, ALDH2 *2-Sham, WT-MI, ALDH2 *2-MI, n=5, 5, 5, and 6, respectively). \**P* < 0.05, \*\**P* < 0.01 and \*\*\**P* < 0.001 from 2-way ANOVA test. Results were reported as the mean ± SEM. **G**, representative immunoblotting images of ferroptosis marker proteins TFRC, ACSL4-S (short time exposure), ACSL4-L (long time exposure), HMOX1, and FTH in the infarct tissues 3-day after AMI or sham group (n=3 for each group). **H**, quantitation of **G**. \**P* < 0.05, \*\**P* < 0.01 and \*\*\**P* < 0.001 from 2-way ANOVA. Results were reported as the mean ± SD. **I**, cardiac mRNA level of Tfrc, Acsl4, Hmox1, and Fth in the infarct tissues 3-day after AMI or sham group (WT-Sham, ALDH2 *2-Sham, WT-MI, ALDH2 *2-MI, n=5, 5, 5, and 6, respectively). \**P* < 0.05, \*\**P* < 0.01 and \*\*\**P* < 0.001 from 2-way ANOVA test. Results were reported as the mean ± SEM. ACSL4, acyl-CoA synthetase long chain family member 4; FTH, ferritin heavy chain 1; HMOX1, heme oxygenase 1; TFRC, transferrin receptor.

Previous studies found that protein levels of ALDH2 *2 were significantly decreased in human and mouse liver tissues, whereas Aldh2 mRNA levels did not significantly change.^42^ We analyzed the ALDH2 protein in mouse heart and found that protein levels of ALDH2 *2 mutant significantly decreased without affecting mRNA levels. Interestingly, we also found that MI led to the upregulation of HIF-1α at both protein and mRNA levels compared to the sham, and this upregulation was significantly attenuated in ALDH2 *2 post-AMI (Figure 3E and 3F). Compared with the WT sham and AMI group, we found that ferroptosis marker proteins TFRC, ACSL4 and HMOX1 upregulated obviously, whereas FTH was downregulated in the mice infarct tissues of ALDH2 *2 at 3-day post-AMI (Figure 3G and 3H). We also detected antioxidative proteins in ferroptosis and found that DHODH was decreased, whereas DHFR was increased after AMI both in WT and ALDH2 *2 infarct tissues. But there was no difference between WT and ALDH2 *2 in neither sham nor AMI group (Figure S3A and S3B). Notably, we observed the changes of enzymatic products of Alox15, 13-HODE in mice serum with ALDH2 *2 variant (Figure 2E), 15-HETE in the mice heart tissues of ALDH2 *2 with Fer-1 treatment (Figure S2F), however, the protein levels in the infarct tissues did not significantly change (Figure S3E). We also checked protein markers of other cell death pathways, such as necroptosis (MLKL), apoptosis (caspase-3 and caspase 8), in the infarct heart tissues. MLKL increased slightly in ALDH2 *2 sham left ventricular tissues but there existed no differ post-AMI compared to WT (Figure S3C and S3E), whereas cleaved caspase 8 (c-Casp 8) increased as Casp-3 decreased in ALDH2 *2 infarct tissues post-AMI compared to WT (Figure S3D and S3F).

Previously work also found Aldh2 mRNA downregulated as p53 mRNA upregulated post-AMI in WT mice.^43^ We further verified that protein levels of TFRC, ACSL4, and p53 were significantly upregulated, whereas ALDH2 *2 was downregulated and GPX4 did not show significant change in the infarct tissues (Figure S3G and S3J). As atrium and brain regions were prone to be affected by ischemia, we only observed upregulation of p53 and p-p53 in the atrium of ALDH2 *2 without affecting TFRC, ACSL4, HMOX1 and GPX4 (Figure S3H and S3K). In the brain, ACSL4 levels were increased accompanied with decreased levels of SLC7A11 and GPX4 in ALDH2 *2 post-AMI (Figure S3I and S3L). Interestingly, ALDH2 protein also downregulated in ALDH2 *2 brain tissues (Figure S3I and S3L). Although TFRC, ACSL4 and HMOX1 protein levels were specifically upregulated in the infarct tissues of ALDH2 *2 compared to WT post-AMI, the mRNA levels of Tfrc, Acsl4, and Hmox1 in ALDH2 *2 infarct tissues showed no increase (Figure 3I).

Taken together, all these data strongly support a role of ferroptosis in exacerbating MI injury in ALDH2 *2 through upregulating protein levels of TFRC, ACSL4, and HMOX1 in the infarct tissues without affecting mRNA levels.

### ALDH2 Deficiency Predisposes Cardiomyocytes to Ferroptosis by Promoting Tfrc/ Acsl4/ Hmox1 mRNA Translation

Next, we set out to investigate the mechanisms leading to the upregulation of proteins critical for ferroptosis without affecting mRNA levels. To do so, we treated human cardiomyocyte AC16 cells with various inducers of cell deaths. And found Staurosporine (apoptosis), Rapamycin (autophagy), H_2_O_2_ (necroptosis), did not appear to make a difference between ALDH2 WT and ALDH2 knockdown (Figure S4A-C). Furthermore, ALDH2 deficiency cells were more sensitive to cell death with RSL3 induced ferroptosis (Figure S4E). We employed this cell model to examine whether protein degradation is involved in the upregulation of ferroptosis proteins in ALDH2 *2 in MI. Next, we tested the protein levels of TFRC, ASCL4, and HMOX1 at various time points with treatment of protein synthesis inhibitor cycloheximide (CHX) in the context of RSL3 incubation. We found that there was no significant difference in the decay of any of these proteins with ALDH2 knockdown (Figure 4A, 4B and S4E). Lysosomal inhibitor BA (Bafilomycin A1) or proteasome inhibitor MG132 co-treatment could rescue part of protein degradation (Figure S4F). All these data strongly suggested that protein degradation pathways play a limited role in the upregulation of TFRC, ACSL4 and HMOX1 proteins in exacerbating MI injury in ALDH2 *2 via ferroptosis.

**Figure 4.**
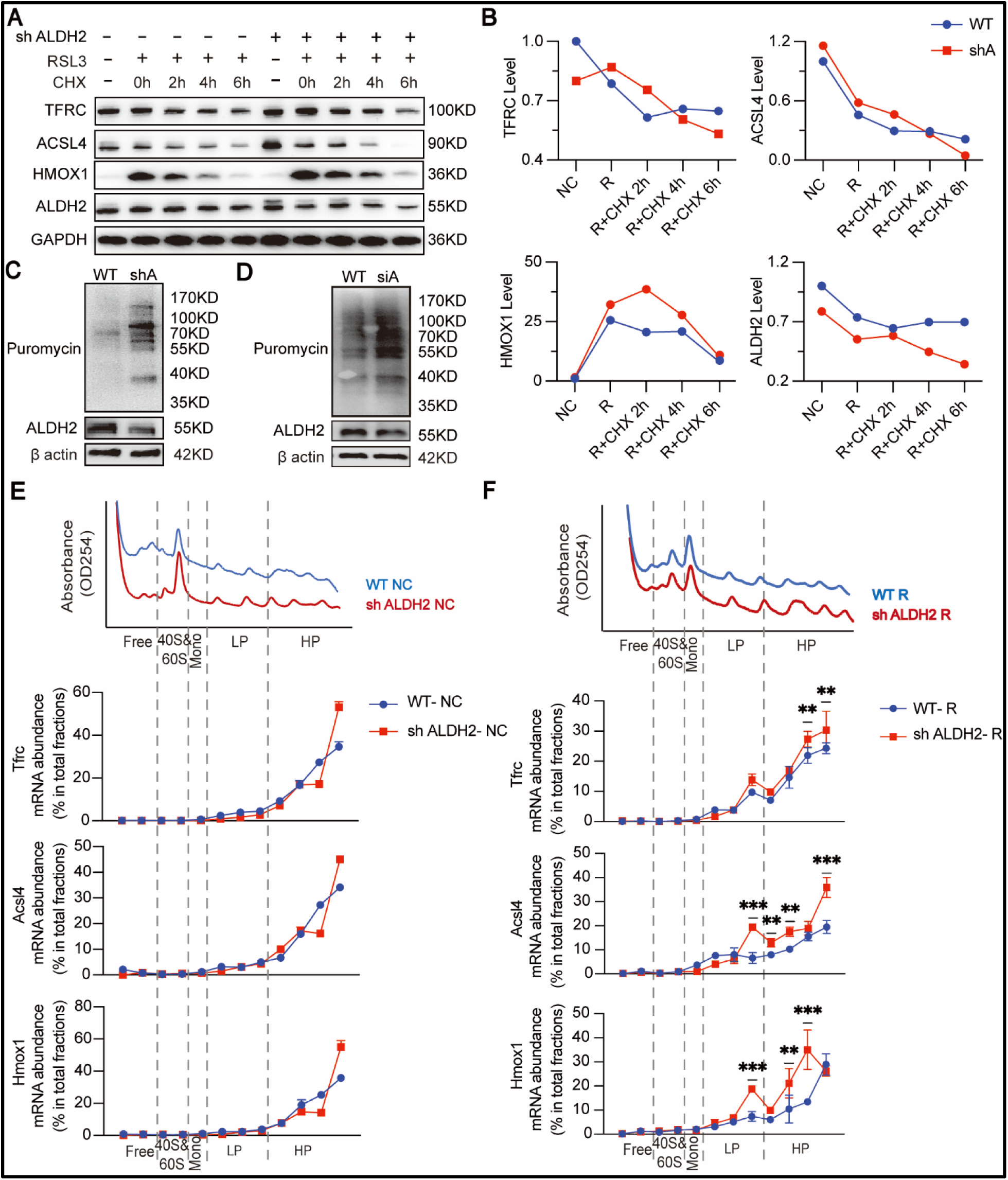
ALDH2 deficiency aggravated cell ferroptosis by promoting Tfrc/ Acsl4/ Hmox1 translation. **A**, 1 μM RSL3 treatment for 12h to induce ferroptosis, then AC16 WT and shALDH2 cells were treated with 5 μg/mL CHX for 0h, 2h, 4h and 6h. Western blot analyzed TFRC, ACSL4, HMOX1 and ALDH2 in the indicated cells. **B**, quantitation of **A**. **C** and **D,** western blot analysis of puromycylated peptides in AC16 sh-NC, shALDH2 and si-NC, siALDH2 cells following treatment with RSL3 12h and puromycin (10 μg/mL) for 10 min prior to cell lysis. **E** and **F**, polysome profiles of NC and shALDH2 in AC16 cells. The isolated fractions were free, 40S&60S, monosome, LP (light polysome), and HP (heavy polysome). RT-qPCR determining the mRNA levels of Tfrc, Acsl4 and Hmox1 enriched in each fraction. Gapdh as the positive control and membrane protein hY1 as the negative control. Results were reported as the mean ± SD. \*\**P* < 0.01 and \*\*\**P* < 0.001 from Student t test. CHX, cycloheximide; NC, normal control; RT-qPCR, real-time polymerase chain reaction.

We further interrogated the role of protein translation in modulating the protein levels by Ribo-Lace, a technique that could specifically analyze active ribosome during translation.^44^ Interestingly, the active ribosome was upregulated in ALDH2 deficiency cells (Figure 4C and 4D). Ribo-seq also found that the codon usage frequency of initiation codon AUG was significantly higher, and the translation efficiency of Tfrc and Acsl4 increased higher. Hmox1 had the increased tendency, whereas Hif-1α decreased significantly in ALDH2 deficiency cells during ferroptosis (Figure S5A and S5B). To verify the translation efficiency of Tfrc, Acsl4 and Hmox1 during ferroptosis, we conducted a polysome profiling experiment. After ribosomes were separated by glucose density gradient solution, free, 40S, 60S, monosome, light polysome (LP), and heavy polysome (HP) were isolated for qPCR detection (Figure 4E, 4F, S6A, and S6B). During ferroptosis, the mRNA levels of Tfrc, Acsl4 and Hmox1 were significantly enriched in high polysome fraction of ALDH2 deficiency cells than in WT (Figure 4F and S6A), whereas in normal cells, the mRNA content did not show significant difference (Figure 4E, S6A, and S6B).

Together, all these data suggest that ALDH2 deficiency makes cardiomyocytes more susceptible for ferroptosis through enhancing translation of TFRC, ACSL4, and HMOX1.

### ALDH2 Physically Interacts with Eukaryotic Initiation Factors 3E (eIF3E)

To study how ALDH2 affects protein translation in ferroptosis, we hypothesized that ALDH2 might directly interact with protein translation machinery to regulate the translation of TFRC, ACSL4, and HMOX1. In a previous study,^28^ we performed proteomic analysis of ALDH2 interaction proteins with ALDH2 and found that eukaryotic initiation factors 3 complex (eIF3) were among the potential interacting proteins (Table S1). eIF3 is one of the most complex initiation factors with 13 different subunits (eIF3a–m) in mammals and has been implicated in many steps of translation initiation.^45, 46^ As a member of eIF3, eIF3E has been implicated in the translation of Tfrc, Acsl4 and Hmox1.^45, 47, 48^ We speculated that decreased ALDH2 proteins may promote the translation of these three ferroptosis-related proteins through eIF3E in the context of ferroptosis. Noteworthy, elF2a and eIF3E did not change between the WT and ALDH2 *2 groups either in the sham or MI, whereas eIF3E were significantly downregulated in the MI groups of both WT and ALDH2 *2 (Figure S7A). But MI did not significantly affect the protein expressions of eIF3E or mitochondrial membrane voltage dependent anion channel 1 (VDAC1) (Figure S7B), suggesting ALDH2 *2 plays no limited role in inducing mitochondria stress post-AMI.

Next, we employed an online tool Gromacs and predicted a potential protein interaction of ALDH2 and elF3E (Figure 5A). To experimentally validate the interaction, we performed co-immunoprecipitation (co-IP) experiments using the left ventricular tissues from three patients with heart transplantation after heart failure (Table S2) and found that ALDH2 and eIF3E were pulled down with each other (Figure 5B). Consistently, ALDH2 and eIF3E were pulled down together in the left ventricular tissues of WT mice and less eIF3E was pulled down by ALDH2 antibody in ALDH2 *2 mice (Figure 5C and 5D). Similar result was also observed in in AC16 cells (Figure S7C). Furthermore, eIF3E co-localized with ALDH2 in the spatial immunofluorescent experiments but not with the mitochondrial membrane protein TOM20, suggesting eIF3E and ALDH2 primarily interact with each other in the cytoplasm (Figure 5E). Since the peptides 1-346 of eIF3E is critical for the translation function, we constructed four plasmids and overexpressed in 293T cells with HA tagged full-length eIF3E, truncated eIF3E (1-346), and Flag tagged rs671(ALDH2 *2)-Flag, and ALDH2-Flag. We found that the eIF3E translation peptide was critical for interacting with ALDH2 (Figure S7D). According to the Gromacs prediction, H12, Q241, T248 and T287 were potential interacting sites. We purified four point-mutant proteins: eIF3E H12A, Q241A, T248A, and T287A (Figure S8A, S8B and S8C), and found that all these four mutants significantly attenuated the interactions between eIF3E and ALDH2 (Figure S8D). Interestingly, the mutant ALDH2 rs671 binds to elF3E to similar extent to the WT ALDH2 (Figure 5F and S8D).

**Figure 5.**
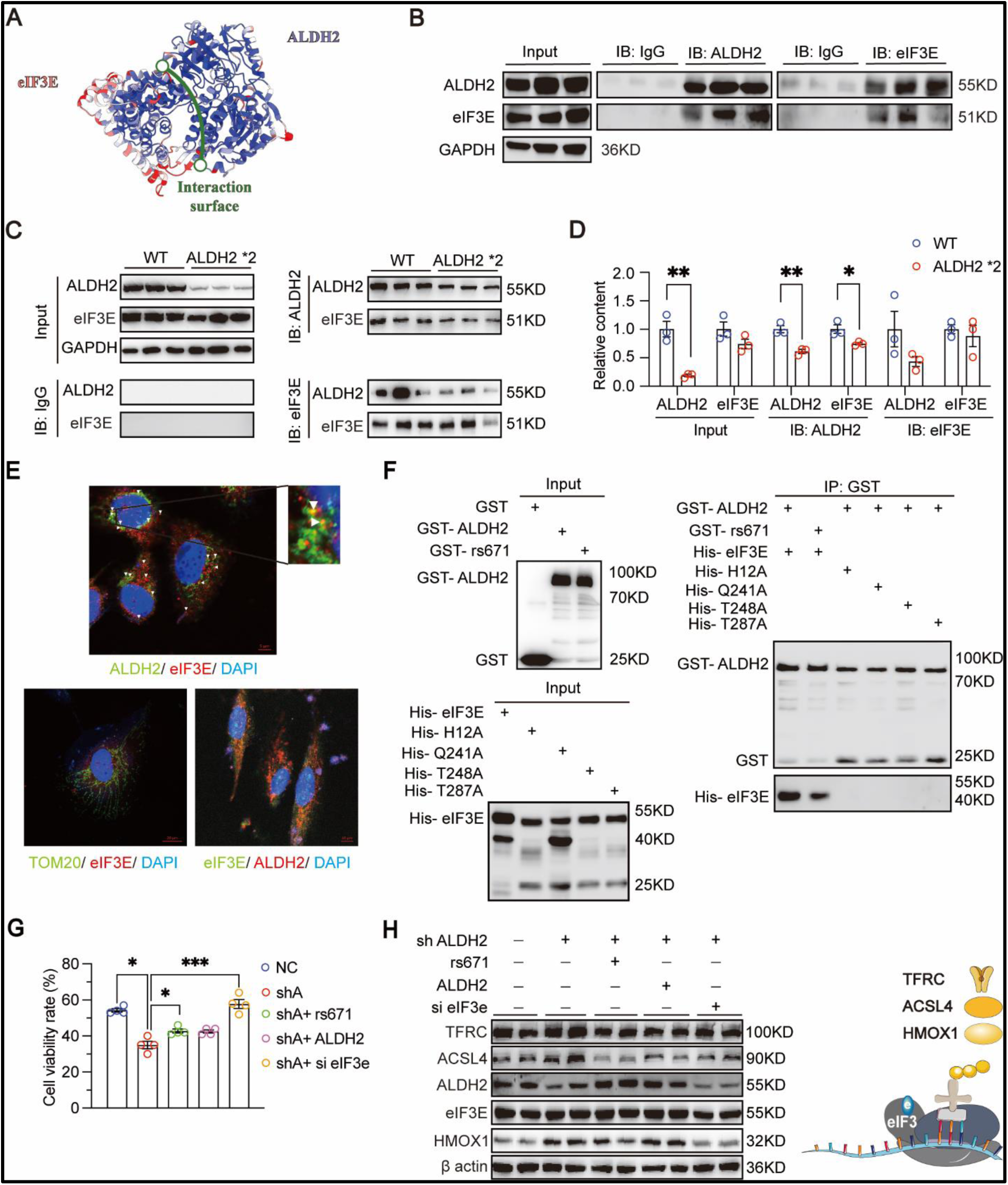
ALDH2 deficiency promotes Tfrc, Acsl4, and Hmox1 translation by direct interaction with eIF3E. **A**, gromacs prediction of the interaction sites between ALDH2 and eIF3E. **B**, co-IP results in the left ventricular tissues from heart failure patients (n=3, respectively). **C**, co-IP results in the left ventricular tissue of mice (n=3 for each group). **D**, quantitation of proteins shown in **C**. Results were reported as the mean ± SD. \**P* < 0.05 and \*\**P* < 0.01 from 2-way ANOVA test. **E**, prepared AC16 WT cells for immunofluorescence co-staining of eIF3E with ALDH2 or TOM20. TOM20 was used as the mitochondrial marker. Scale bar, 5 to 20μm. **F**, recombinant His-tagged eIF3E (WT), His-tagged eIF3E (H12A), His-tagged eIF3E (Q241A), His-tagged eIF3E (T248A), and His-tagged eIF3E (T287A) were incubated with GST-tagged ALDH2 or GST-tagged ALDH2 rs671 mutant in GST pull-down assay. **G**, cell viability detection following 1 μM RSL3 treated 12h. Results were reported as the mean ± SD. \**P* < 0.05 and \*\*\**P* < 0.001 from Student t test. **H**, western blot analysis of TFRC, ACSL4, ALDH2, eIF3E, and HMOX1 protein contents among the five genotype cells after RSL3 treatment. Schematic diagram depicting the eIF3E modulates TFRC, ACSL4, and HMOX1 translation. co-IP, coimmunoprecipitation; eIF3E, eukaryotic initiation factors 3E; GST, glutathione S-transferase.

Together, these results suggested that ALDH2 directly interacts with eIF3E in the cytoplasm to affect the protein translation. Although both WT ALDH2 and ALDH2 *2 bind to eIF3E, the decreased protein levels in ALDH2 *2 is primarily responsible for upregulating ferroptosis related proteins, including TFRC, ACSL4, and HMOX1, in the context of AMI.

### Cardiomyocyte-specific Knockdown of EIF3E Improves ALDH2 *2 Heart Function by Decreasing Ferroptosis in MI

To provide further evidence that ALDH2/eIF3E interaction is important for regulating ferroptosis through the protein translations of key proteins in ferroptosis in the context of MI, we knocked down eIF3E expression in AC16 cells and found that knockdown eIF3E completely rescued cell survival in the context of ferroptosis and ALDH2 deficiency, whereas overexpression of ALDH2 or ALDH2 rs671 partially rescued cell survival (Figure 5G). The levels of lipid peroxides increased significantly in ALDH2 deficiency cells, but overexpression of ALDH2 rs671 mutant (ALDH2 *2) or ALDH2 WT protein attenuated the levels of lipid peroxides (Figure S9A), consistent with the cell survival rate detected by Mito Tracker (Mito633) (Figure S9B and 5G). Moreover, GSH peroxidase enzyme activity was significantly increased in the ALDH2 deficiency cells, whereas the GPX activity was decreased with overexpression of ALDH2 rs671 mutant or ALDH2 WT (Figure S9C and S9D). Lipidomic analysis also found increased levels of oxidized phospholipids in the context of ferroptosis and ALDH2 deficiency, which were significantly decreased with overexpression of ALDH2 WT, rs671, and silencing eIF3E (Figure S9E). Consistently, protein levels of TFRC, ACSL4 and HMOX1 were significantly decreased with eIF3E knockdown during ferroptosis and ALDH2 deficiency (Figure 5H). Interestingly, the mRNA level of Tfrc, Acsl4 did not significantly change with or without RSL3 treatment (Figure S9F and S9G). Collectively, all these data indicated that eIF3E is critical in regulating ferroptosis induced by ALDH2 deficiency.

To further verify ALDH2 *2 aggravates cardiomyocyte ferroptosis through eIF3E during AMI, we employed MI mouse model with cardiomyocyte-specific eIF3E knocked down (KD) (Figure 6A). Cardiomyocyte-specific eIF3E knockdown decreased scar area significantly only in ALDH2 *2 mice but not in WT mice 28-day post-AMI (Figure 6B). Furthermore, the cardiac functions of LVEF, LVFS and LVIDs were also significantly improved only in ALDH2 *2 mice with cardiomyocyte-specific eIF3E knockdown (Figure 6C, 6D and S10A). It is noteworthy that cardiomyocyte-specific eIF3E knockdown did not lead to cardiomegaly and myocardial hypertrophy after MI as indicated by the levels of WGA (wheat germ agglutinin) and Ki67 (cell proliferation) staining on heart sections harvested at 28-day after MI (Figure 6E and 6F). Consistently, the increased LDH activity and levels of oxidized phospholipids in ALDH2 *2 infarct tissues at 3-day post-AMI were reduced by cardiomyocyte-specific eIF3E knockdown (Figure 6G and 6H). The Fe^2+^ levels were also reduced in ALDH2 *2 infarct tissues compared to WT mice with eIF3E cardiomyocytes-specific knockdown (Figure 6I). eIF3E was significantly reduced at both mRNA and protein levels in the infarct tissues of ALDH2 *2 cardiomyocyte-specific eIF3E knockdown mice (Figure 6J, and S10B-D). We further found that eIF3E cardiomyocytes-specific knockdown significantly reduced the TFRC, ACSL4, and HMOX1 protein content caused by ALDH2 *2 mutant but had no effect on ALDH2 protein level (Figure 6J, and S10B-D). We did not observe significant difference of the mRNA levels of Tfrc, Acsl4, and Hmox1 in the infarct tissues between WT and ALDH2 *2 post-AMI (Figure S10D). Moreover, previous studies found that Mineralocorticoid receptor (MR) plays a key role in crosstalk between liver and heart in promoting cardiac repair post MI.^49^ Interestingly, MR was significantly reduced after cardiac-specific eIF3E knockdown, which warrants future study (Figure S10D).

**Figure 6.**
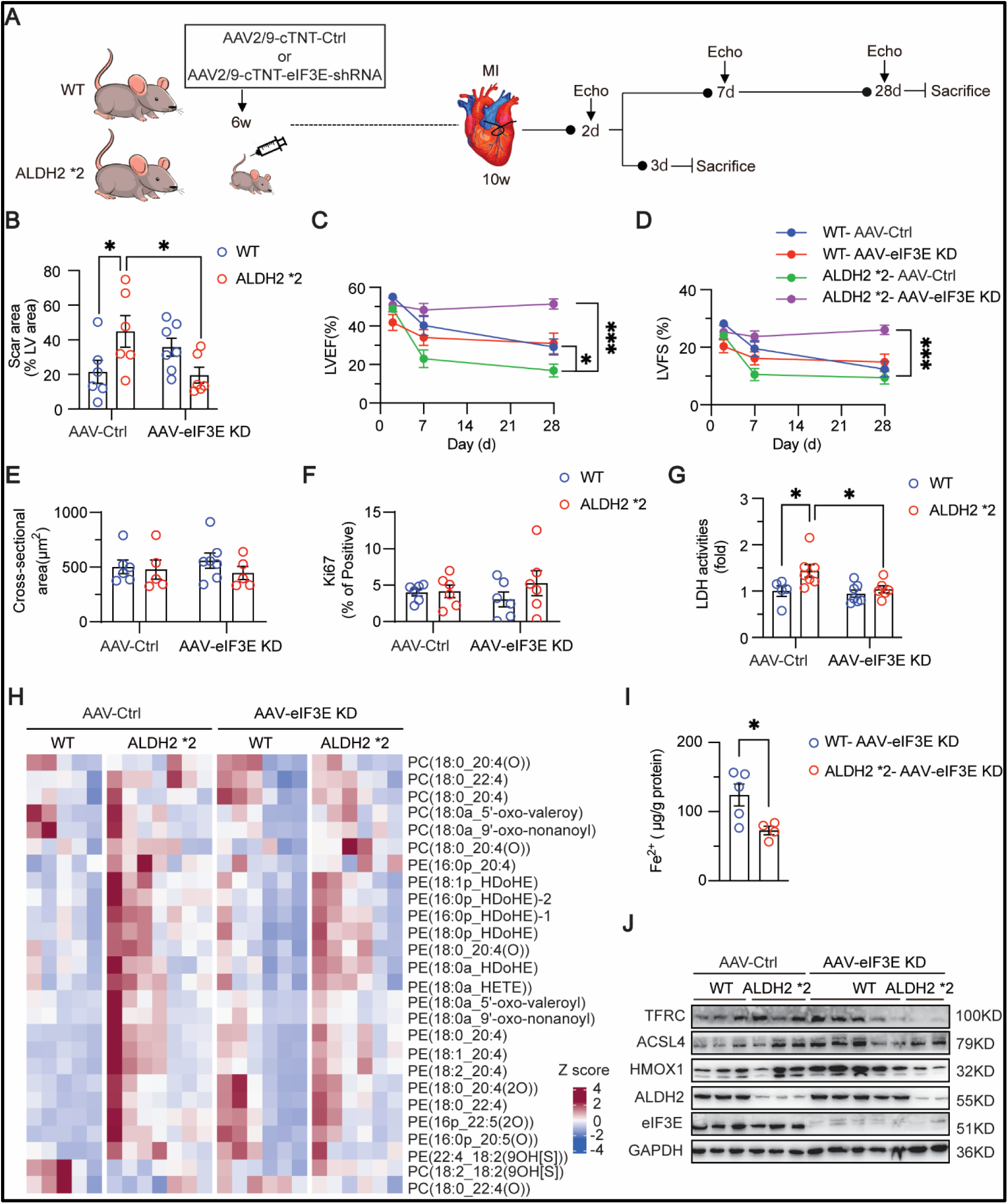
Cardiomyocyte-specific knockdown of eIF3E improves ALDH2*2 heart function by decreasing ferroptosis. **A**, schematic diagram depicting the experimental strategy for WT and ALDH2 *2 mice. 6 weeks old mice were injected with AAV2/9-cTNT-control (Ctrl) or AAV2/9-cTNT-eIF3E-shRNA virus that has been verified in cells through IV. **B**, cardiac scar areas analyzed 28-day after MI (WT-AAV-Ctrl, ALDH2 *2-AAV-Ctrl, WT-AAV-eIF3E KD, and ALDH2 *2-AAV-eIF3E KD, n=6, 6, 7, and 6, respectively). Results were reported as the mean ± SEM. \**P* < 0.05 from Student t test. **C** and **D**, cardiac functions measured in mice, including LVEF and LVFS. Results were reported as the mean ± SEM. \**P* < 0.05 and \*\*\**P* < 0.001 from 2-way ANOVA test. **E** and **F**, quantification of **E**, WGA, and **F**, Ki67 staining to assess cardiomyocyte sectional area in the indicated groups 28 days after MI. Results were reported as the mean ± SEM. **G**, fold changes of LDH activity in the serum at day 3 of AMI (n=5, 7, 7, and 6, respectively). Results were reported as the mean ± SEM. *P < 0.05 from 2-way ANOVA test. **H**, corresponding oxidized phospholipids in the infarct tissues of mice 3-day post-AMI. **I**, the contents of Fe (II) in the infarct tissues of WT and ALDH2 *2 with cardiomyocyte-specific knockdown of eIF3E pretreatment 3-day post-AMI. Results were reported as the mean ± SEM. \**P* < 0.05 from Student t test. **J**, TFRC, ACSL4, HMOX1, ALDH2, and eIF3E protein level in the infarct tissues 3-day post-AMI. Results was reported as the mean ± SEM. \**P* < 0.05, \*\**P* < 0.01, and \*\*\**P* < 0.001 from 2-way ANOVA test. Echo, echocardiography; IV, intravenous injections; KD, knockdown; WGA, wheat germ agglutinin.

Taken together, these findings suggest that targeting cardiomyocyte eIF3E might be a potential therapeutic strategy in protecting against cardiac injury through inhibiting ferroptosis during AMI for ALDH2 *2 carriers.

## DISCUSSION

Substantial amount of clinical and experimental evidence has implicated ferroptosis in the MI but the underlying mechanisms remains poorly defined. Although lipid peroxidation is a core hallmark of ferroptosis, the roles of ALDH2 in ferroptosis in CVD are much less understood.^17, 29^ In this study, we provided strong evidence that ferroptosis is involved in patients and mice carrying ALDH2 *2 variant. Mechanistically, we found that ALDH2 affects ferroptosis through directly interacted with eIF3E to modulate translation of critical proteins involved in ferroptosis, including TFRC, ACSL4, and HMOX1, whereas decreased protein levels of ALDH2 in ALDH2 *2 mutant predispose cardiomyocytes to ferroptosis through upregulating the translation of these proteins (Figure 7). Furthermore, knocking down elF3E or inhibiting ferroptosis by Fer-1 effectively restored cardiac function during ischemic acute heart failure for ALDH2 *2 mice.

**Figure 7.**
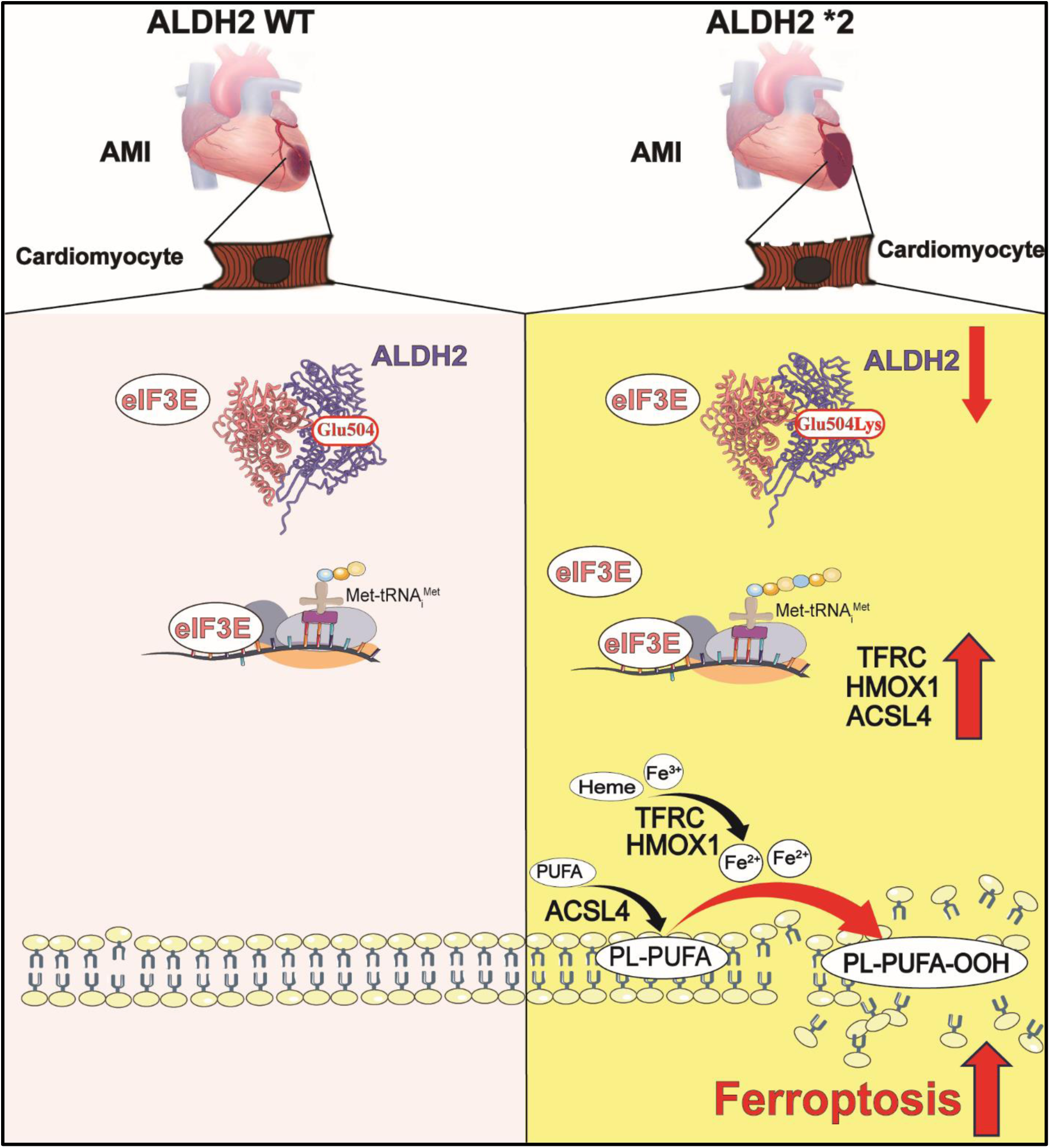
ALDH2 mutation in ALDH2*2 predisposes cardiac cells to ferroptosis in heart failure: ALDH2 *2 facilitates ferroptosis-related protein translation through interaction with eIF3E to promote heart failure post AMI. Left, ALDH2 WT heart post-AMI. Right, ALDH2 *2 heart post-AMI. Red lines represent the promoted process in ALDH2 *2 mutant post-AMI. ALDH2 *2, aldehyde dehydrogenase 2 rs671 mutant. eIF3E, eukaryotic initiation factors 3e. PUFA, polyunsaturated fatty acid. PL-PUFA, PUFA-containing phospholipids.

Human clinical studies have linked ALDH2 *2 mutant with increased risks of CVD, especially in the context of MI, but the underlying mechanism remain poorly defined.^17^ A majority of previous reports attributed the increased risks of CVD to the accumulation of alcohol-derived acetaldehydes or lipid aldehydes generated from lipid peroxidation, such as 4-HNE,^9^ due to the decreased enzyme activity of ALDH2 in this loss-of-function variant.^29^ Consistently, activation of ALDH2 enzymatic activity, such as Alda-1, has been explored for the treatment of MI, especially for ALDH2 *2 carriers.^17, 50^ Furthermore, nitroglycerin tolerance was a result of ALDH2 inactivation, and ALDH2 activation by Alda-1 may protect patients with MI from nitroglycerin-induced cardiac injury while maintaining the cardiac benefits of the increased nitric oxide concentrations produced by nitroglycerin.^51^ A recent study identified ovarian tumor (OUT) deubiquitinase 5 (OTUD5) as a novel protector against 4-HNE triggered ferroptosis in MI/IR injury, whereas ALDH2 knockdown aggravates myocardial ferroptosis due the accumulation of 4-HNE.^30^ In a separate study, ALDH2 deficiency exacerbates MI injury through neutrophil extracellular trap, which can be prevented by leukotriene C4 inhibition.^23^ Interestingly, emerging evidence showed that ALDH2 *2 has decreased protein levels in various tissues although the mechanisms remain unknown.^16, 52–54^ Conceivably, decreased ALDH2 enzymatic functions in ALDH2 *2 can be either due to ALDH2 mutation or decreased ALDH2 levels, and several experimental studies have linked decreased ALDH2 enzyme function to heart failure with excessive alcohol intake.^17, 55^ However, we and others have shown that prevalence of alcohol drinking was obviously lower in ALDH2 *2 carriers in heart failure patients (Table 1), suggesting unknown mechanisms underlying increased CVD risks in an alcohol-independent mannner.^56^ Our previous studies uncovered multiple non-enzymatic roles of ALDH2 in increased risks of atherosclerosis, including hepatic HDL biogenesis, cholesterol synthesis, and macrophage foam cell formation and efferocytosis.^17, 26–28, 57^ ALDH2*2 upregulates *de novo* cholesterol synthesis by stabilizing HMGCR, the rate-limiting enzyme in cholesterol synthesis and the target of statins.^27^ Interaction of ALDH2 with LDLR and AMPK is important in macrophage foam cell formation.^26^ ALDH2 *2 polymorphism increased the ubiquitination of RAC2 and promoted its degradation, thereby having an opposite role in necrotic core formation.^58^

In this study, we identified a novel mechanism by which ALDH2 interacts with protein translation machinery elF3E to affect the protein translations of critical proteins in ferroptosis. This study represents the first example for the critical role of protein translation in ferroptosis in MI although previous studies have demonstrated that ALDH2 is involved in various forms of cardiomyocyte deaths, such as autophagy, necroptosis, apoptosis, and shown to protect against ischemia-reperfusion injury.^55, 59–61^ Interestingly, a previous study identified a noncanonical function of eIF4E in limiting the mitochondrial protein ALDH1B1 in detoxifying 4-HNE and increasing the susceptibility to ferroptosis in cancer cells.^62^ In our study, we observed upregulation of ferroptosis related protein TFRC, ACSL4, and HMOX1 specifically in the infarct area of ALDH2 *2 mice without affecting mRNA levels. Our previous study have found the translation initiation factors eIF3E was in the list of proteins enriched by ALDH2 antibody in liver cells.^28^ In addition, Tfrc, Hmox1, and Acsl4 were on the list of mRNAs affected by eIF3E.^48^ The protein interaction of ALDH2 with eIF3E was further supported by co-IP experiments and co-localization by immuo-fluorescence. Lastly, our present study has not only provided substantial evidence for the involvement of ferroptosis in MI for ALDH2 *2 mutant carriers, but also implied that cardiomyocyte ferroptosis plays an important role in the early stage of MI in mice (2 to 3-day after MI).

Although a body of evidence has linked ferroptosis in MI, the research field has been hampered by the lack of proper biomarkers for ferroptosis in human clinical studies.^9, 10^ In this study, we developed targeted lipidomic methods to measure the levels of lipid oxidation products and bioactive lipids derived from common PUFAs, such as linoleic acid and arachidonic acid.^63^ These state-of-the art techniques have been regarded as the gold standard to measure oxidative stress-induced lipid peroxidation *in vivo* in the context of ferroptosis.^11, 63^ We found that the levels of lipid peroxidation products increased significantly in ALDH2 *2 mice infarct tissues, ALDH2 deficient AC16 cells, and in the plasma of ALDH2 *2 heart failure patients. Furthermore, several arachidonic acids metabolites and arachidonic acid increased obviously in the plasma of ALDH2 *2 heart failure patients and in the serum of ALDH2 *2 mice 3-day post-AMI. 14,15-DHET/14,15-EET ratio, a strongest predictor of 1-year death, increased significantly in ALDH2 *2 AMI patients. A previous studies have found that the CoQ10 and BH4 are cardioprotective.^37, 64, 65^ Intriguingly, however, CoQ10 and BH4 significantly decreased in the plasma of patients with acute heart failure. Future research is warranted to investigate the mechanisms how CoQ10 and BH4 are downregulated in ALDH2 *2 acute heart failure patients post-MI.

## CONCLUSION

In summary, our study demonstrates that ferroptosis is involved in cardiomyocyte deaths and worsen heart failure in ALDH2 *2 patients post-AMI. Mechanistically, ALDH2 interacts with protein translation factor eIF3E to modulate translation of critical proteins in ferroptosis, whereas ALDH2 *2 also predispose cardiomyocytes to ferroptosis through upregulating TFRC, ACSL4, and HMOX1. Inhibition ferroptosis plays a protective role by inhibiting lipid peroxides and ferrous accumulation in ALDH2 *2. Targeting the ALDH2 *2/ eIF3E axis may be an effectively strategy for restoring cardiac function and treating heart failure induced by myocardial infarction for ALDH2 *2 carriers.

## Data Availability

All data for this manuscript will be available upon request

## ARITICLE INFORMATION

### Acknowledgments

We thank the mass spectrometry platform, molecular biology/ biochemistry/cell technology platform, experimental animal platform, and biological sample pathology analysis platform at Shanghai Institute of Nutrition and Health (SINH), Chinese Academy of Sciences (CAS).

### Sources of Funding

This research was funded by grants from and National Natural Science Foundation of China (32241017 and 32030053), and the National Key Research and Development Program of China (2022YFC2503300), Shenzhen Medical Research Fund (SMRF B2302042), Science and Technology Commission of Shanghai Municipality (22140903300), and RGC General Research Fund (9043653), startup funds from the City University of Hong Kong (9380154 and 7006046), RGC Theme-based Research Scheme (8770011), and TBSC Project fund and Futian research project (9609327). DWY was supported by Shenzhen Clinical Research Center for Metabolic Diseases (Shenzhen Science, Technology and Innovation [2021]287), Shenzhen Center for Diabetes Control and Prevention (NO: SZMHC [2020]46).

### Disclosures

None.

## Nonstandard Abbreviations and Acronyms

AA: arachidonic acid
ACSL4: acyl-coA synthetase long chain family member 4
AHF: acute heart failure
ALDH2: acetaldehyde dehydrogenase 2
AMI: acute myocardial infarction
BH4: tetrahydrobiopterin
BNP: brain natriuretic peptide
CoQ10: Coenzyme Q10
CRP: C-reactive protein
CVD: cardiovascular disease
DHET: dihydroxyeicosatrienoic
Echo: echocardiogram
EET: epoxy eicosatetraenoic acid
EF: ejection fraction
EIF3E: eukaryotic translation initiation factor 3 subunit E
Fer-1: ferrostatin-1
FS: fraction shortening
HF: heart failure
HMOX1: heme oxygenase 1
KD: knockdown
LC-MS: liquid chromatography-mass spectrometry
LDH: lactate dehydrogenase
LV: left ventricle
MI: myocardial infarction
NC: normal control
PC: glycerophosphatidylcholine
PE: glycerophosphatidylethanolamine
TFRC: transferrin receptor

## Notes

### Competing Interest Statement

The authors have declared no competing interest.

### Author Declarations

The study protocol was approved by the institutional ethics committee of Changhai Hospital of Shanghai on research in humans.

